# A minimal model for household-based testing and tracing in epidemics

**DOI:** 10.1101/2020.10.29.20222406

**Authors:** Greg Huber, Mason Kamb, Kyle Kawagoe, Lucy M. Li, Aaron McGeever, Jonathan Miller, Boris Veytsman, Dan Zigmond

## Abstract

In a previous work [1], we discussed virus transmission dynamics modified by a uniform clustering of contacts in the population: close contacts within households and more distant contacts between households. In this paper, we discuss testing and tracing in such a stratified population. We propose a minimal tracing strategy consisting of random testing of the entire population plus full testing of the households of those persons found positive. We provide estimates of testing frequency for this strategy to work.

## 1. Introduction

Widespread COVID-19 epidemics have made lockdown and shelter-in-place policies common in many countries. These policies make the simplifying panmictic assumption of classic epidemiological models less appropriate than during the early pandemic phase [2, 3, 4]. Lockdown and shelter-in-place policies make the contrasting assumption relevant: the population is divided into small groups (“households”) with a high proportion of contacts within the household and relatively lower number of contacts between households. An understanding of epidemics dynamics in such an inhomogeneous population is important to predict the spread of infection and to design measures to mitigate and suppress it.

In a previous work [1] we proposed a minimal model describing epidemics in such a compartmentalized population. The key feature of the model was the separation of infection into a fast intra-household mode and a slow inter-household mode. We showed that the model predicts a non-trivial dependence of the reproduction number R0 on the household size *H*: linear in *H* for relatively small households, and as 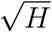 for larger households. We argued that the insights from the minimal model are important to understand the dynamics of the epidemics and the effect of different policies, even while much more detailed models exist [5, 6, 7, 8, 9].

In the present paper, we discuss a policy for testing and tracing in the above model. It is well established that testing and tracing are among the best tools for suppression of epidemics [10]. The household model provides a ready analog of this policy: if a household member is known to have the disease, we quarantine the entire household (or, in a less strict version of the policy, we test everyone and quarantine any member who tests positive).

This model can describe several situations. First, we can randomly test representatives of different households, triggering entire household testing if the representative is positive. In another situation, household testing is triggered by a household member exhibiting symptoms of the disease. In the latter case, we assume most infected persons to be asymptomatic or mildly symptomatic [11], thus the presence of a highly symptomatic individual points to other infections in the household.

An attractive feature of this model is the low tracing effort required and the minimal degree of compliance necessary: we simply trace the members of the household known to contain infected persons. If this measure is accompanied by some degree of inter-household tracing, the suppression efforts would be even more successful.

## 2. Mean-field approach

The assumptions made are close to those in [1]. An individual can be in one of the following states (we assume no natural immunity): susceptible *s*, infected *i*, recovered *r* and quarantined *q*, as shown in Figure 1. Of course, a quarantined individual also changes their state to recovered (or dead), but since we are interested only in the spreading of the infection, that change does not influence our model. We assume that the recovery rate (or, in a more pessimistic interpretation, recovery and death rate) is described by the parameter *γ*. The rate of discovery and quarantine is described by the parameter *κ*.

**Figure 1.**
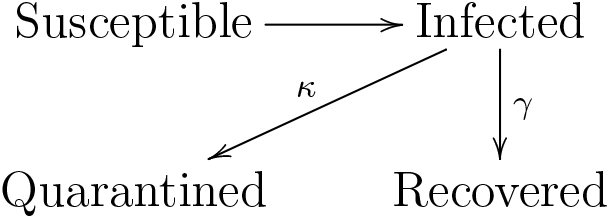
State diagram for an individual.

A household state can therefore be described by a four-dimensional vector of integers, (*s, i, q, r*). We make the same assumption as in [1]: the intra-household rate of infection is fast. Then the simplified state diagram for a household is shown in Figure 2. The mean-field transition rate from state *G* (everybody is infected) to state *Q* (everybody is quarantined) is determined in the following way. Consider a household with *H* infected members. The probability for any of them to exhibit symptoms or test positive during time interval *dt* is *κ dt*. Therefore the probability that no one in the household becomes symptomatic is (1 − *κ dt*)^*H*^, and the probability that someone becomes symptomatic is 1 − (1 − *κ dt*)^*H*^ = *κH dt*.

**Figure 2.**
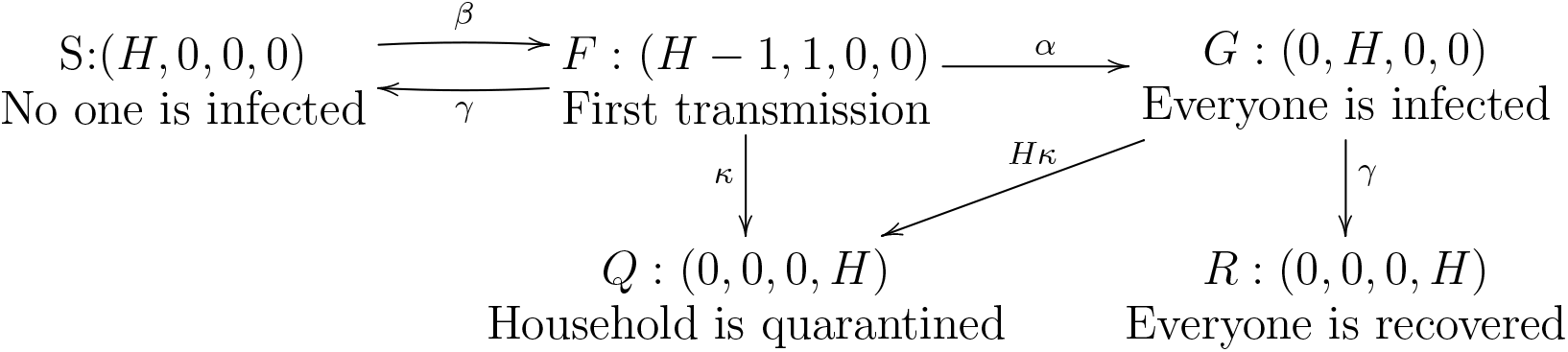
Simplified state diagram for a household.

The recovery rate for a household where everyone is infected is the same as that for an individual. A fully infected household has a state vector (0, *H*, 0, 0) and becomes (0, 0, 0, *H*) upon recovery. A household with one infected individual has a state (*H* − 1, 1, 0, 0) and becomes (*H* − 1, 0, 0, 1) after that individual recovers. Since the following analysis only looks at short time scales, the final state after individual recovery (and, for that matter, full household recovery) won’t play a role in the dynamics. So, in Figure 2, we choose to draw a recovery arrow from (*H* − 1, 1, 0, 0) to (*H*, 0, 0, 0). This choice has two advantages: it avoids a proliferation of relevant states *i*.*e*., the number of possible states does not grow with *H*) and the number of susceptibles in the population is nearly the same, even for times beyond the initial regime.

Following [1], let *F* be the number of non-quarantined households with just one person infected, *G* be the number of non-quarantined households with everyone infected, and *N* be the total number of persons. Let *α* be the intra-household infection rate and *β* the inter-household infection rate. Then, in the limit of fast intra-household transmission, we get:

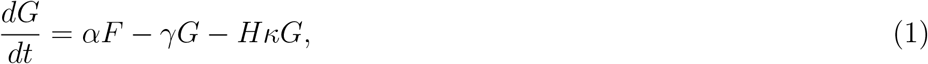

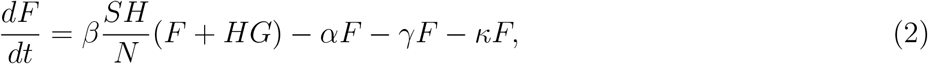

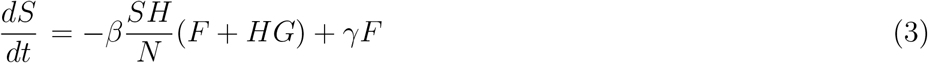

We are interested in the initial regime, when *SH ≈ N*. The largest eigenvalue is then 2

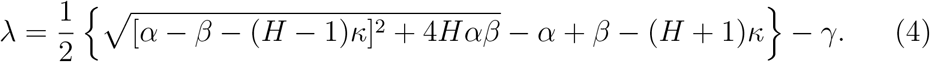

The outbreak is localized if *λ* < 0. Let us designate by *κ* = *κ*_0_ the threshold testing rate at which *λ* = 0.

Let us assume that the intra-household transmission rate *α* exceeds both the inter-household transmission rate *β* and the testing rate *κ*. Expanding equation (4) for large *α*, we get

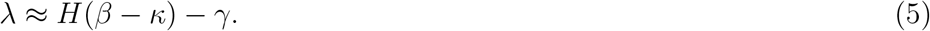

This result does not depend on the rate of intra-household transmission *α*, which is reasonable since that is effectively infinite in this approximation. The minimal testing rate necessary to stop the epidemics is then

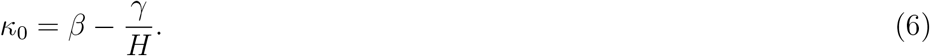

One consequence of this result is apparent: in the limit of large households, we need to test everybody at a rate *κ* exceeding the individual transmission rate *β*.

For finite rates *α*, equation (4) can be used to calculate the values of *κ* for which epidemics do not spread. This is done in Figure 3. From this figure, one can see that, in large households, the threshold value for testing rate is approximately *β* for all reasonable values of intra-house transmission rate. In the following sections, we discuss the changes to these predictions when a more general approach than mean-field theory is adopted.

**Figure 3.**
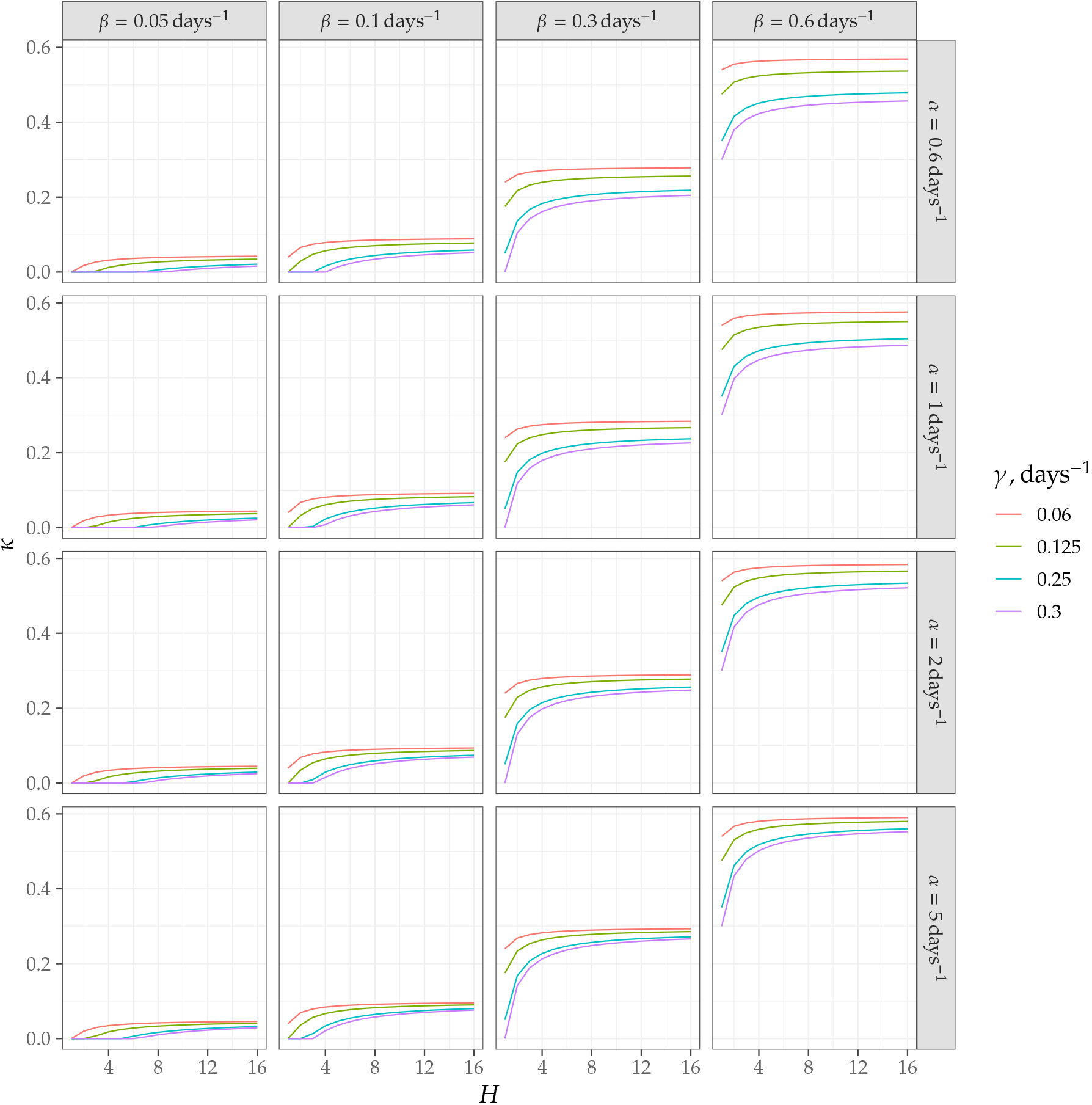
Threshold values of testing rates *κ* needed to stop the epidemics in the household model.

## 3. Numerical modeling

We performed numerical simulations to validate the analytical model of household infections described in the above section. In each scenario that we simulated, we imagined a population of *N* = 200 000 individuals and divided these into households of a specific size. We then initialized a random subset of 20 households with one infected individual. For each step, we divided the infection into four phases:

i. *Panmictic phase*: each actively infected individual can infect any uninfected individual in the simulation, with a probability 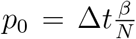, where *β* is the daily rate of infection in the panmictic phase, *N* is the population size and Δ*t* is the time step of the simulation. We assume that all new infections from this step are found in previously uninfected households.
ii. *Household phase*: each household with a single infection transitions to a fully infected state with a probability Δ*tα*, where *α* is the rate parameter for in-householdinfection.
iii. *Testing phase*: infected individuals are discovered with a probability *κ*Δ*t* in single-infection households, and *Hκ*Δ*t* in fully-infected households. If an infected individual is detected inside a household, that household is removed from the panmictic phase of the infection, corresponding to a quarantine.
iv. *Recovery phase*: Infected households can recover fully with a probability *γ*Δ, at which point they are removed from the panmictic phase of the simulation and incur no more new infections

We ran simulations for Δ*t* = 0.1 days and various combinations of *α, β, γ* and *H*. The results for *γ* = 0.125 days^−1^ and *κ* = 0.06 days^−1^ are shown in Figure 4 together with mean field predictions based on numerical solution of equations (1), (2), and (3) without the assuming *SH ≈ N* (*i*.*e*. both inside and outside the linear regime). The figure shows that the outbreak is indeed localized at *β* < *κ* and spreads at *κ* much smaller than *β*. The dynamics at *β* close to *κ* depends on the intra-household transmission rate *α* (assumed to be infinite in the analytical approximation). Also, as expected, the discrepancy between mean-field theory and simulations is largest at the phase boundary, *β ≈ κ*.

**Figure 4.**
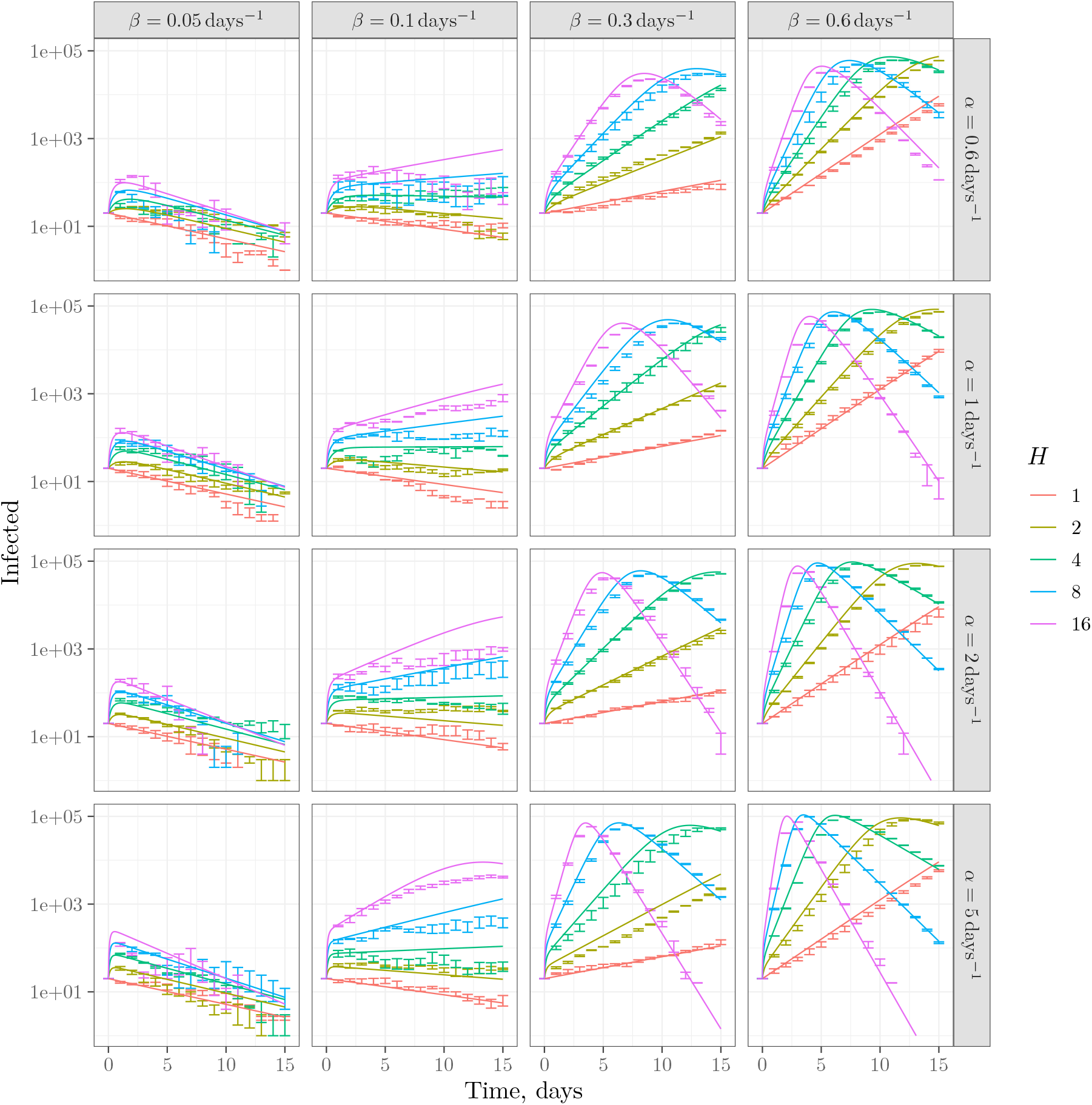
Simulations and mean field predictions for the testing and tracing model for *γ* = 0.125 days^−1^ and *κ* = 0.06 days^−1^.

The total number of infected cases during the epidemics is shown in Figures 5–7. The figures show that testing and (limited-to-a-household) tracing indeed can stop epidemics.

**Figure 5.**
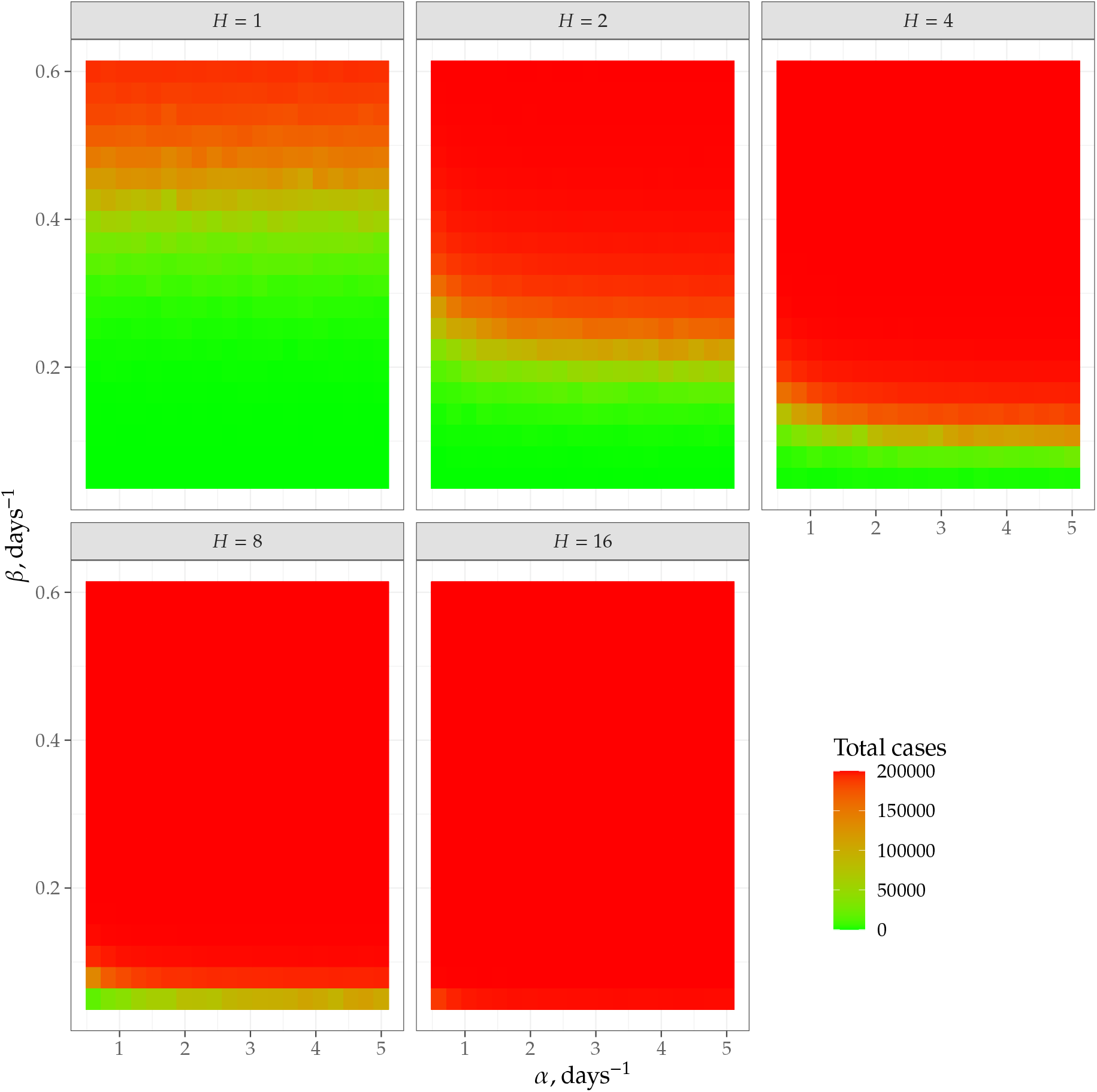
Total number of infected cases (out of 200000) for the no-testing scenario.

## 4. Testing and discovery of infected patients

The parameter *κ* in our model is the true rate of discovery of infected persons. We should stress that this is not the rate of testing: to get the rate of discovery we need to multiply the testing rate by the sensitivity of the test. For the case of COVID-19, testing technology is under rapid development [12], but already the sensitivity and specificity of swab tests with PCR have been widely discussed in the literature [13, 14, 15, 16]. Specificity seems, with general agreement, to exceed 95% [15]. Sensitivity of a single test in practice is believed to vary depending on the conditions in the field under which testing is performed and because standardization is inconsistent [14]. A commonly used value for single-test sensitivity is 70%. The data indicate that this value varies widely over the duration of infection and the reliability of this number is uncertain [13].

Recently, it has been observed that the emphasis on sensitivity of a one-time test may not always be justified. Multiple lower-sensitivity (and, therefore, potentially less expensive and technically demanding) tests administered at suitable intervals may be more effective at preventing spread of infection. This observation shifts attention away from the analytical sensitivity of a one-time test (for example), toward the sensitivity of a testing regimen [12]. The perfect-sensitivity testing scheme proposed here represents such a regimen, folding neatly into the recent observations [12], with a prescription for obtaining an inter-test interval with a defined impact on the spread of infection. It may be worth understanding how relatively low-sensitivity testing affects the dynamics of the model studied here.

## 5. Discussion and conclusions

The two parts of the phrase “testing and tracing” are, in a sense, complementary. If we test everyone, we do not need tracing contacts to suppress epidemics. On the other hand, perfect tracing would obviate the need for testing. A real strategy combines testing and tracing to compensate for deficiencies in either: we randomly test because we do not have perfect tracing, and we trace contacts because we cannot test everyone frequently enough. In this paper we discussed the situation where a minimal version of tracing is implemented: we trace only the most frequent contacts of an infected person, *i*.*e*. the people sharing a household with them. Obviously, any tracing strategy must include these contacts, so this is indeed the *minimal* tracing policy, as discussed in the Introduction.

The main conclusion of our analysis is that the minimal approach works as long as the testing rate is high enough. The minimal testing rate (up to corrections of the order 1*/H, H* being the household size) is described by a simple and intuitively acceptable rule: the time between the tests for a randomly selected individual should be smaller than the average time to infect someone during inter-household contacts. This rule can be used to estimate the required testing frequency – or to determine whether inter-household tracing is necessary with said testing frequency.

The mean-field theory result (equation (6)) suggests that increasing household size is always detrimental to the control of epidemics. However, our simulations reveal that for aggressive testing the increase of household size may actually help to control the epidemics (compare the last panels on Figure 7). The reason for this is the following: in our mean-field approximation, we assumed a very fast spread of infection within households, so that one infected individual quickly gives rise to *H* infected individuals, which, in turn, infect other households. In actuality, there is some time (of the order of 1*/α*) during which the number of infected is lower than *H*. If any of these infected individuals is tested, we may stop the spread within the household. The average time between a member of the household being tested is 1*/*(*Hκ*), so this effect is noticeable at *H ≈ α/κ*, which agrees well with Figures 5–7.

**Figure 6.**
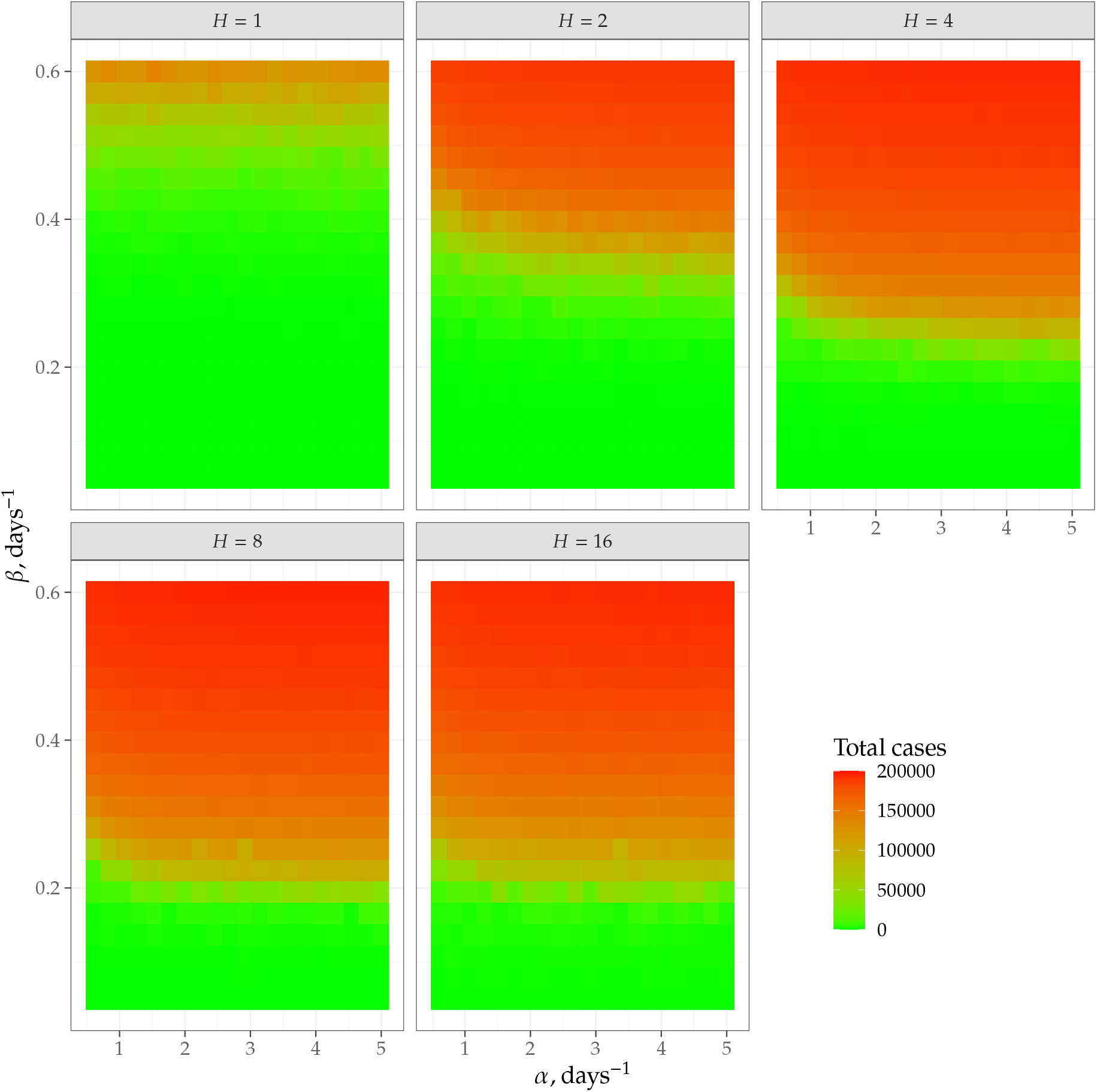
Total number of infected cases (out of 200000) for a moderate-testing scenario, *κ* = 0.126 days^−1^.

**Figure 7.**
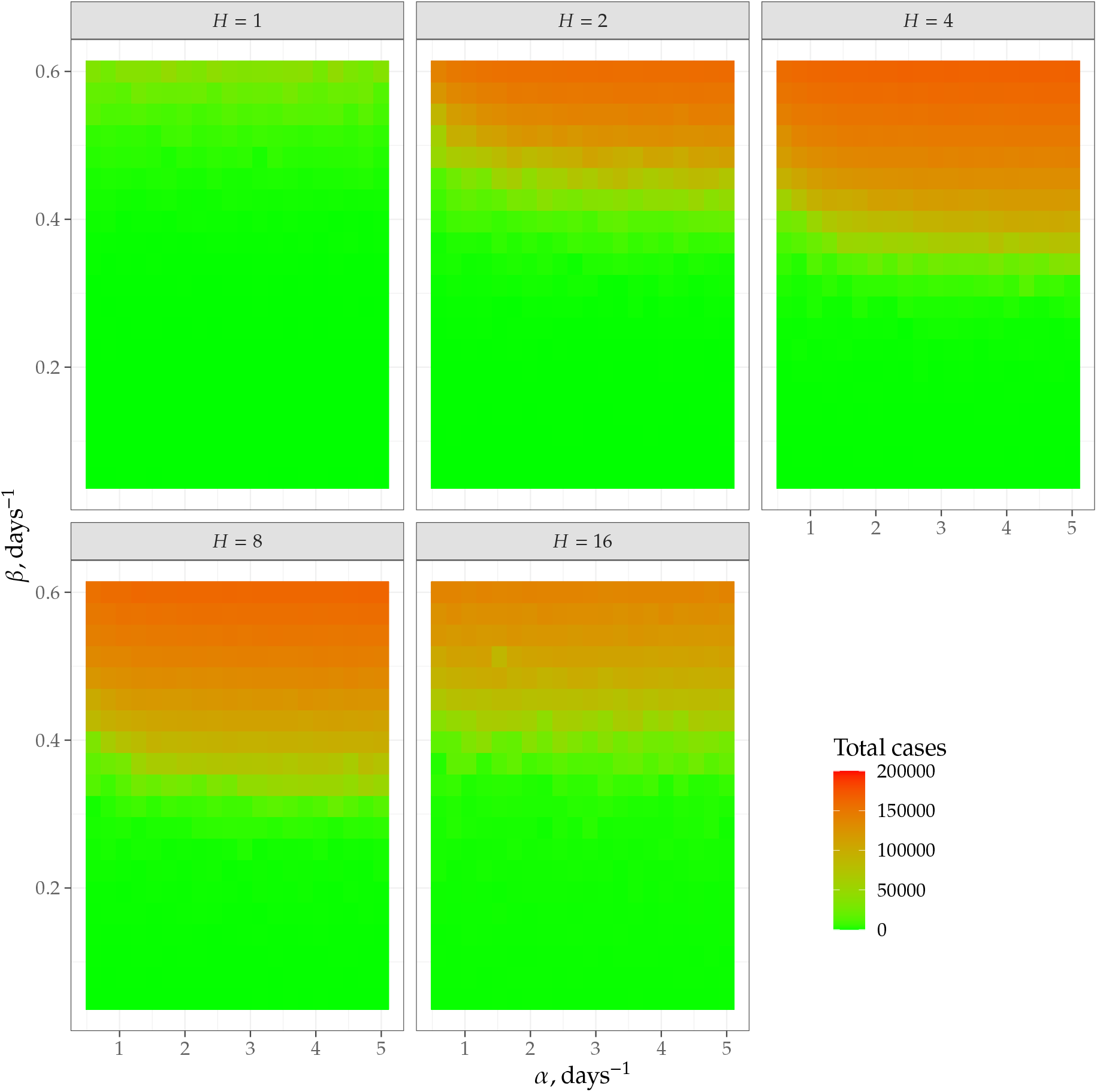
Total number of infected cases (out of 200000) for a high-testing scenario, *κ* = 0.221 days^−1^.

As for the real world application of these results, one should keep in mind several points. First, as discussed in Section 4, the testing rate should be increased for low-sensitivity tests, keeping the discovery rate high enough. Second, studies have shown that socioeconomic factors impact the ability to quarantine, which means many symptomatic people will be unable to isolate even with a positive test. In this sense, the calculations here provide lower estimates for the required levels of testing.

## 6. Data and code availability

The code and data for this paper are available at https://github.com/Kambm/HouseholdDynamics.

## Data Availability

The data and code are available

https://github.com/Kambm/HouseholdDynamics

